# AI-Powered Effective Lens Position Prediction Improves the Accuracy of Existing Lens Formulas

**DOI:** 10.1101/2020.10.29.20222539

**Authors:** Tingyang Li, Joshua D. Stein, Nambi Nallasamy

## Abstract

**Aims:** To assess whether incorporating a machine learning (ML) method for accurate prediction of postoperative anterior chamber depth (ACD) improves the refraction prediction performance of existing intraocular lens (IOL) calculation formulas.

**Methods:** A dataset of 4806 cataract patients were gathered at the Kellogg Eye Center, University of Michigan, and split into a training set (80% of patients, 5761 eyes) and a testing set (20% of patients, 961 eyes). A previously developed ML-based method was used to predict the postoperative ACD based on preoperative biometry. This ML-based postoperative ACD was integrated into new effective lens position (ELP) predictions using regression models to rescale the ML output for each of four existing formulas (Haigis, Hoffer Q, Holladay, and SRK/T). The performance of the formulas with ML-modified ELP was compared using a testing dataset. Performance was measured by the mean absolute error (MAE) in refraction prediction.

**Results:** When the ELP was replaced with a linear combination of the original ELP and the ML-predicted ELP, the MAEs ± SD (in Diopters) in the testing set were: 0.356 ± 0.329 for Haigis, 0.352 ± 0.319 for Hoffer Q, 0.371 ± 0.336 for Holladay, and 0.361 ± 0.331 for SRK/T which were significantly lower than those of the original formulas: 0.373 ± 0.328 for Haigis, 0.408 ± 0.337 for Hoffer Q, 0.384 ± 0.341 for Holladay, and 0.394 ± 0.351 for SRK/T.

**Conclusion:** Using a more accurately predicted postoperative ACD significantly improves the prediction accuracy of four existing IOL power formulas.

## INTRODUCTION

The estimation of postoperative intraocular lens position is essential to intraocular lens power calculations for cataract surgery. Norrby and Olsen have reported that inaccuracy in the prediction of the postoperative anterior chamber depth (ACD) is the number one source of error for postoperative refraction prediction.[1,2] In addition to its vital role in intraocular lens (IOL) formulas, the postoperative ACD is also a critical variable in ray tracing, where the uncertainty in the postoperative ACD directly affects the accuracy of the results. Methods to improve the accuracy of the prediction of postoperative ACD have been studied for decades. In first-generation formulas, the lens position was represented by a constant. Later, more and more preoperative biometric variables such as the axial length and the corneal power were added to calculate the postoperative IOL position. In 1993, Holladay first proposed the name “effective lens position (ELP)” to indicate the location of the lens as it relates to a given optical model of the eye.[3] Although the ELP was constructed to estimate the position of the IOL, practically the ELPs calculated using existing formulas (e.g., SRK/T) are not accurate estimates of the physical location of the IOL.[1,4] This is mainly because the ELPs in those formulas were formulated to account for different formula-specific assumptions and regression results.[1] In view of the limitations of the ELP in existing formulas, recently, more efforts have been devoted to constructing ELPs that better reflect the true location of the IOL.[5–9] New IOL power prediction methods have also been developed based on the new-generation ELP prediction methods, and they have shown that using a more accurately predicted IOL position helps to improve the IOL power prediction accuracy.[5]

It is so far largely unexplored whether inserting a more accurately predicted ELP into existing formulas improves refraction prediction accuracy. This is an important question because: (1) it provides a fast and efficient way to modify and improve on existing IOL formulas whose reliability has been tested extensively. (2) such research can provide supports for translating the continued improvements in accuracy in postoperative ACD prediction into better refraction predictions in published formulas. Several previous studies had modified the ELPs in existing formulas in order to achieve better refraction prediction results in certain cataract cases. Modification of ELP calculation in the Haigis formula for sulcus-implanted IOLs was reported to improve performance.[10] Kim et al. adjusted the ELP estimation in SRK/T formulas with the corneal height in post-refractive patients and achieved satisfactory accuracy.[11] It remains to be explored whether improvement of ELP estimates for in-the-bag IOL placement can improve IOL power calculations of existing formulas for general cataract patients.

Since most recently published IOL formulas (e.g., Barrett Universal II[12,13], Holladay 2, Olsen formula[14]) are either not disclosed to the public or do not have the option to customize the value of ELP during the prediction of postoperative refraction, here we applied our previously developed postoperative ACD prediction methods to a dataset of 4806 cataract surgery patients and replaced the ELP estimates in 4 existing IOL formulas: Haigis, Hoffer Q, Holladay, and SRK/T. We combined our machine learning (ML) prediction of true postoperative ACD with the original ELP estimated by each formula and substituted this updated ELP prediction for each formula. We then compared the refraction prediction performance of each formula using its original and enhanced ELP estimates. The findings reported here demonstrate that existing formulas can benefit from improved methods for predicting true postoperative ACD.

## MATERIALS AND METHODS

### Postoperative ACD prediction machine learning model

In previous work,[15] we developed a machine learning-based postoperative anterior chamber depth (ACD) prediction model, which predicts the postoperative anterior chamber depth (in mm) based on preoperative biometry. Here in the presented study, an ACD prediction machine learning model was trained using the method and dataset (847 patients, 4137 eyes) described in the previous research. The dataset was composed of the preoperative and postoperative biometry measured by the Lenstar LS900 optical biometers (Haag-Streit USA Inc, EyeSuite software version i9.1.0.0) at the University of Michigan’s Kellogg Eye Center. The postoperative ACD was defined as the distance from the front surface of the cornea to the front surface of the intraocular lens (IOL). The postoperative ACD predicted by the machine learning model is referred to as *ELP*_*ML*_ in this manuscript.

### Data collection

In this study, biometry records were collected using the same approach as for the development of the ML postoperative ACD prediction model.[15] The inclusion criteria were: (1) patients who had cataract surgery (CPT = 66984 or 66982) but no prior refractive surgery and no additional surgical procedures at the time of cataract surgery. (2) the implanted lens was an Alcon SN60WF single-piece acrylic monofocal lens (Alcon, USA). Each case in the dataset corresponds to one operation of a single eye with preoperative and postoperative information. The preoperative information includes the measurements of the axial length (AL), lens thickness (LT), anterior chamber depth (ACD), flat keratometry (K1), steep keratometry (K2), and the average keratometry which was calculated as 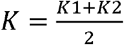. The postoperative information includes the postoperative refraction (spherical component SC and cylindrical component CC) where the time when it was recorded was closest to one month (30 days) after surgery. Since the patients were measured in a lane of 10 feet long (3.048 meters), which was shorter than the standard length of 20 feet (6 meters), the SC was adjusted for the vergence distance by adding 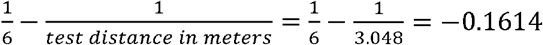 according to Simpson and Charman’s recommendation.[16] The spherical equivalent (SE) refraction was therefore calculated as *SE refraction = (SC* − 0.1614) + 0.5*CC*. Samples that were used to train the postoperative ACD prediction machine learning model were excluded from the dataset so that the dataset better simulates unseen samples.

The dataset in total consisted of 4806 patients (**Figure 1**). The dataset was split into a training dataset used for the development of the methods and a testing dataset used for performance comparison. 80% of the patients were randomly assigned to the training set, and the rest of the patients (20%) were assigned to the testing set. For patients who had more than one associated case in the testing set (i.e., patients who had both eyes operated on), one case was randomly selected to ensure each patient had the same weight when the prediction performance was evaluated. At the end of this process, the training set had 3845 patients (5761 eyes), and the testing set had 961 patients (961 eyes).

**Figure 1.**
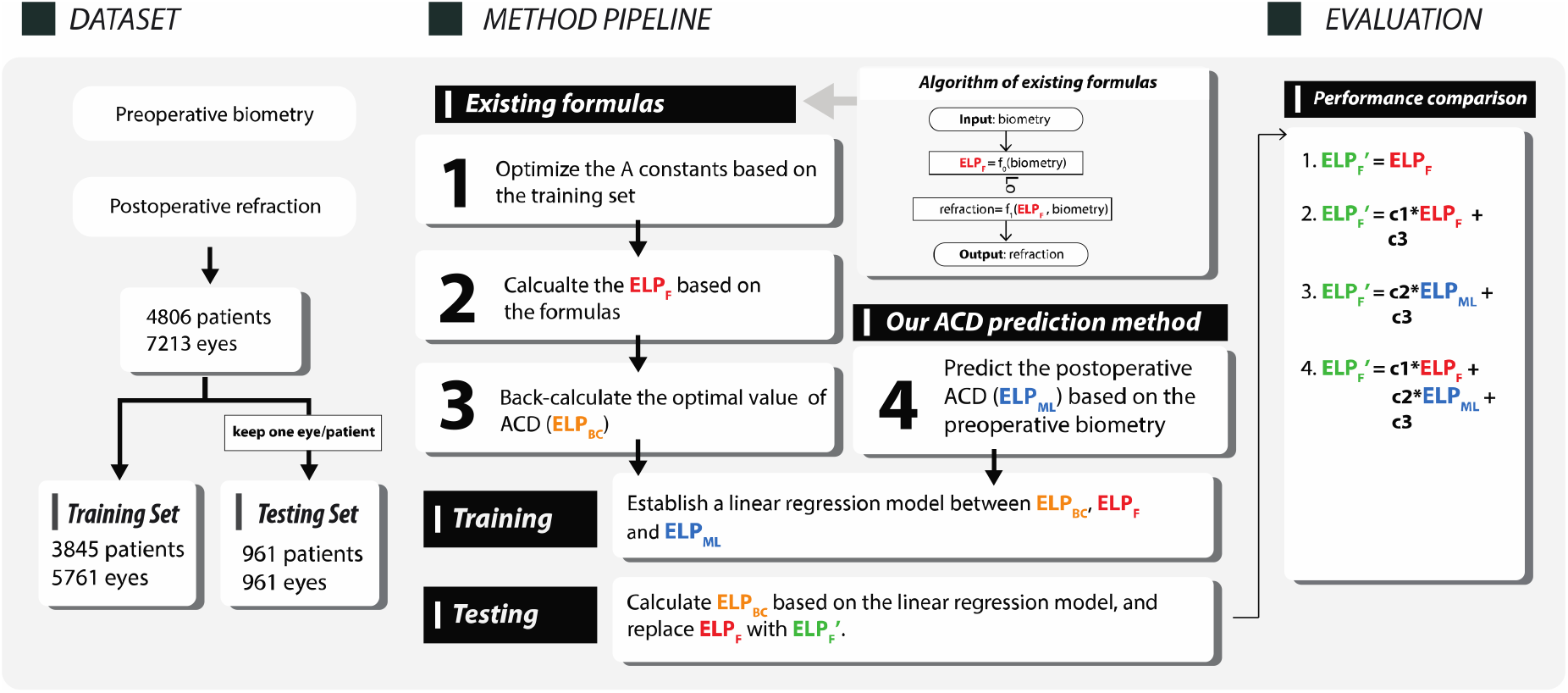
The analysis pipeline of the presented study. *ELP*_*F*_ = the effective lens position (ELP) estimated by the existing formulas. *ELP*_*ML*_ = the postoperative anterior chamber depth (ACD) predicted by the machine learning method. *ELP*_*BC*_ = the back-calculated ELP (see main text). 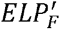 is a term that refers to a new ELP that is used to replace the *ELP*_*F*_ in the existing formulas.

### Linear regression model

We implemented four existing formulas (Haigis, Hoffer Q, Holladay, and SRK/T) in Python based on their publications.[17–24] The existing formulas calculated the effective lens position (*ELP*_*F*_) as a function of the preoperative biometry (**Figure 1**): *ELP*_*F*_ *= f*_0_*(biometry)*. The predicted ELP (*ELP*_*F*_) was then used to predict the postoperative refraction: *refraction = f*_1_*(ELP*_*F*_, *biometry)*. Here, the goal was to reduce the refraction prediction error by replacing *ELP*_*F*_ with a different value, 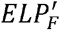. Our approach involves two steps: (1) finding the theoretically most optimal ELP values, (2) modeling the most optimal ELP with *ELP*_*F*_ and the ML-predicted postoperative ACD, denoted *ELP*_*ML*_.

In the first step, the most optimal ELP (denoted *ELP*_*BC*_) was found by the standard method of back-calculating the ELP when the predicted refraction was set to equal the true refraction (i.e., *f*_1_ *(ELP*_*BC*_, *biometry) = true refraction*). In other words, when 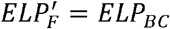, the refraction prediction errors of all patients equal zero. More details on the computation of *ELP*_*BC*_ can be found in **Supplementary materials**.

After the computation of *ELP*_*F*_, *ELP*_*ML*_, and *ELP*_*BC*_, we modeled *ELP*_*BC*_ using a linear function of *ELP*_*F*_ and/or *ELP*_*ML*_ so as to obtain an approximation of the most optimal ELP using available variables. We compared four different approaches of approximating *ELP*_*BC*_ : (1) Original, 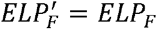 : using the original *ELP*_*F*_, (2) Formula LR, 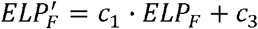 : using linearly adjusted *ELP*_*F*_, (3) ML LR, 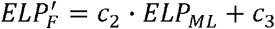: using linearly adjusted *ELP*_*F*_, (4) Formula & ML LR, 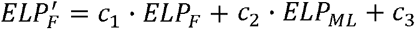: using a linear combination of *ELP*_*F*_ and *ELP*_*ML*_. Here *c*_1_, *c*_2_, and *c*_3_ are constants. Outliers with large refraction errors (i.e., *error* ≥ *mean error + 2 · standard deviation* or *error* ≥ *mean error* − *2 · standard deviation*) were excluded for each formula before establishing the linear regression model, in order to obtain better modeling results. The refraction prediction errors were calculated as *error = predicted refraction* − *true refraction*. The linear regression was performed using scikit-learn 0.20.3.

On the testing set, 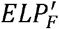 was calculated based on the values of *c*_1_, *c*_2_, and *c*_3_ obtained through linear regression. The predicted refraction was calculated as *refraction = f*_1_*(ELP*_*F*_ *′, biometry)*. The mean absolute error (MAE), median absolute error (MedAE) and mean error (ME) were calculated for performance comparison.

### A-constant optimization

The A-constants for the formulas were optimized based on the training dataset so that the mean error in refraction prediction was closest to zero. The A-constants were optimized separately for the unmodified formulas and formulas with a modified ELP estimate (see **Supplementary Materials**). The optimized A-constants for the original formulas were: a0 = −0.733, a1 = −0.234, a2 = 0.217 for Haigis, ACD constant = 5.724 for Hoffer Q, surgeon factor = 1.864 for Holladay, and A = 119.089 for SRK/T (**Table S1**).

### Statistical analysis

Linear regression analysis was used to assess the significance of the correlation between *ELP*_*F*_, *ELP*_*ML*_, and *ELP*_*BC*_. To test whether the MAE and ME of different methods were significantly different, a Friedman test followed by a post hoc paired Wilcoxon signed-rank test with Bonferroni correction was used. Statistical significance was defined as the p-value <0.05. All the above analyses were performed with Python 3.7.3.

## RESULTS

### Dataset overview

The cases in the training and testing datasets had a similar distribution according to the summary statistics shown in **Table 1**. As elaborated in **Materials and Methods**, we calculated *ELP*_*F*_, *ELP*_*ML*_, and *ELP*_*BC*_ based on the formulas and their optimized A-constants. The mean and standard deviation of the ELPs calculated based on the original formulas were summarized in **Table S2**. *ELP*_*BC*_ and *ELP*_*F*_ had similar mean values in contrast to *ELP*_*ML*_.

**Table 1.** The summary statistics for the patient demographics for the training and testing dataset. For the age at surgery, preoperative biometry, and postoperative refraction, the mean ± SD (standard deviation) is shown in the table.

| Characteristic | Training set | Testing set |
| --- | --- | --- |
| **Gender** | Male: 2514 eyes (43.6%),<br>Female: 3247 eyes (56.4%) | Male: 425 eyes(44.2%),<br>Female: 536 eyes (55.8%) |
| **Age at surgery (years)** | 70.99 ± 9.61 | 70.10 ± 10.24 |
| **Preoperative K (D)** | 43.85 ± 1.64 | 43.90 ± 1.66 |
| **Preoperative AL (mm)** | 24.19 ± 1.40 | 24.20 ± 1.41 |
| **Preoperative LT (mm)** | 4.54 ± 0.45 | 4.53 ± 0.45 |
| **Preoperative ACD (mm)** | 3.24 ± 0.41 | 3.26 ± 0.41 |
| **Postoperative refraction (D)** | -0.53 ± 0.96 | -0.57 ± 0.90 |

The Pearson correlation coefficients (*R*) between *ELP*_*F*_, *ELP*_*ML*_, and *ELP*_*BC*_ were shown in **Table 2**. Three ELP-related variables were positively intercorrelated with each other. The correlation coefficients, *R*, between *ELP*_*BC*_ and *ELP*_*ML*_ were the weakest among the three pairs of variables across all formulas.

**Table 2.** The Pearson correlation coefficients (*R*) between *ELP*_*F*_, *ELP*_*ML*_, and *ELP*_*BC*_. The *ELP*_*BC*_ and *ELP*_*F*_ were calculated using the A constants optimized based on the original formulas. P-values of all correlations were < 0.05. The corresponding scatter plots are shown in **Figure S1**. All *R* were rounded to three decimal places.

| Index | Variable Pairs | Haigis | Hoffer Q | Holladay1 | SRK/T |
| --- | --- | --- | --- | --- | --- |
| **1** | $ELP_F$ vs. $ELP_{ML}$ | 0.751 | 0.676 | 0.698 | 0.636 |
| **2** | $ELP_{BC}$ vs. $ELP_F$ | 0.621 | 0.730 | 0.622 | 0.633 |
| **3** | $ELP_{BC}$ vs. $ELP_{ML}$ | 0.532 | 0.544 | 0.534 | 0.524 |

### Linear regression results on the training set

Linear regression models were established based on the training set and the *R*^2^ of alternative linear models were shown in **Table 3**. The coefficients of the fitted linear regression line are shown in **Table S3**. The mean and SD of the 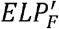 resulting from different models are shown in **Table S4**. For “Formula LR”, the *R*^2^ was larger than that of “ML LR” for all four formulas. For “Formula & ML LR”, the *R*^2^ was larger than that when one of *ELP*_*F*_ and *ELP*_*ML*_ was excluded from the linear combination for all four formulas.

**Table 3.** The *R*^2^ of alternative least-squares linear regression models in the training set. The outlier cases were removed before calculating the above values. The largest *R*^2^ among three methods is marked in bold for each formula. P-values of all correlations were < 0.05.

| Index | Methods | Haigis | Hoffer Q | Holladay1 | SRK/T |
| --- | --- | --- | --- | --- | --- |
| **1** | **Formula LR** | 0.377 | 0.541 | 0.579 | 0.394 |
| **2** | **ML LR** | 0.376 | 0.442 | 0.426 | 0.378 |
| **3** | **Formula & ML LR** | **0.425** | **0.622** | **0.605** | **0.482** |

### Refraction prediction performance comparison on the testing set

We tested the performance of four scenarios on the testing set and summarized the MAE and SD **Table 4**. The mean error (ME) and median absolute error (MedAE) were shown in **Table S5 and Table S6**. Statistical tests were used to compare the difference in the MAEs of different models (see **Materials and Methods**). Using a linear combination of *ELP*_*F*_ and *ELP*_*ML*_, the refraction prediction results of four existing formulas were significantly improved compared to original *ELP*_*F*_ (statistical test results shown in **Table S7** and **Table S8**).

**Table 4.** Performance in the testing set. The mean absolute error (MAE) ± standard deviation (SD) and the percentage reduction in MAE compared to “Original” for alternative linear models in the testing set. All MAE and SD were rounded to three decimal places. The percentage reduction was calculated as 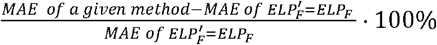. All percentage reduction values were rounded to one decimal place. The method with the smallest MAE among four alternative methods is marked in bold for each formula.

| Index | Methods | Haigis | Hoffer Q | Holladay1 | SRK/T |
| --- | --- | --- | --- | --- | --- |
| **1** | **Original** | 0.373 ± 0.328 | 0.408 ± 0.337 | 0.384 ± 0.341 | 0.394 ± 0.351 |
| **2** | **Formula LR** | 0.373 ± 0.328<br>(0.0%) | 0.374 ± 0.321 (8.3%) | 0.388 ± 0.342 (-1.1%) | 0.391 ± 0.345 (0.8%) |
| **3** | **ML LR** | 0.391 ± 0.346 (-4.8%) | 0.454 ± 0.375 (-21.4%) | 0.434 ± 0.364 (-13.0%) | 0.397 ± 0.344 (-1.5%) |
| **4** | **Formula & ML LR** | **0.356 ± 0.329 (9.0%)** | **0.352 ± 0.319 (22.5%)** | **0.371 ± 0.336 (3.4%)** | **0.361 ± 0.331 (9.1%)** |

We further compared the MAEs of “Original” and “Formula & ML LR” among patients with short, medium, and long axial length (**Table S9**). It was observed that the short and medium axial length groups had a higher percentage decrease in MAE than the long axial length group for Hoffer Q and SRK/T. For Haigis, the medium AL group achieved higher decrease than the other two groups. And for Holladay, the long AL group achieved more decrease in MAE than the other two groups.

## DISCUSSION

In this study, we applied a previously developed machine learning method for postoperative anterior chamber depth (ACD) prediction to an unseen dataset of 4806 cataract surgery patients to assess whether it was possible to improve the performance of existing IOL formulas (Haigis, Hoffer Q, Holladay, and SRK/T) by replacing each formula’s ELP estimate.

We computed three ELP-related quantities: the machine learning-predicted postoperative ACD (*ELP*_*ML*_), formula-predicted ELP (*ELP*_*F*_), and a back-calculated ELP (*ELP*_*BC*_) that minimized the refraction error for each eye in the dataset. They are strongly correlated with each other (**Table 2**), which indicates that (1) *ELP*_*F*_ and *ELP*_*ML*_ are both predictive of the most optimal ELP *ELP*_*BC*_, (2) *ELP*_*F*_ and *ELP*_*ML*_ contain partially overlapping information, which is consistent with our expectation. *ELP*_*ML*_ is an estimation of the value of the true postoperative ACD. On the other hand, the *ELP*_*F*_ was designed by the originators of each formula to serve a similar purpose but was based on the theoretical assumptions in each formula. Our findings are consistent with observations of previous studies that the ELP estimates made by IOL formulas were numerically different from the true postoperative ACD.[9]

Using a training dataset of 3845 patients, we sought to evaluate whether the machine-predicted postoperative ACD, *ELP*_*ML*_, was able to provide information that could be used to refine each formula’s predicted ELP, *ELP*_*F*_. We established regression models between the *ELP*_*ML*_, *ELP*_*F*_, and *ELP*_*BC*_ to evaluate whether a linear combination of *ELP*_*ML*_ and *ELP*_*F*_ used in place of the original *ELP*_*F*_ could lower the refraction prediction error. Using the modified ELPs, we obtained significantly lower mean absolute errors (MAE) in refraction prediction compared to the formulas with the original ELPs on the unseen testing set (**Table 4**). Notably, the accurately predicted postoperative ACD (*ELP*_*ML*_) alone did not outperform the original ELP (*ELP*_*F*_) when it was inserted into the formulas (**Table 4**, row 3 compared to row 1). This is likely because the original method of calculating ELP in each formula compensates for its particular model of the eye and its associated assumptions. Our *ELP*_*ML*_, however, does not have any components that compensate for the assumptions and constants in the formulas. On the other hand, *ELP*_*ML*_ has information about the true postoperative ACD, which it appears can beneficially alter the original ELP estimate.

In this study, the A-constants were optimized separately when *ELP*_*F*_ was replaced with different 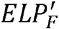. The means of 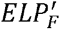, as shown in **Table S4** were numerically close to those of *ELP*_*F*_ as shown in **Table S2**. However, in our method, the similarity between 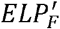 and *ELP*_*F*_ was not among the restrictions and goals of the optimization. The reason that 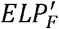 and the original *ELP*_*F*_ have similar means might be that the other parts of each formula put restrictions on the values of ELP in order to obtain reasonable results. This could also be the reason why *ELP*_*BC*_ and *ELP*_*F*_ had similar means as shown in **Table S2**.

The presented method of replacing ELP estimates provides a simple way of improving the prediction performance of existing formulas. While it would be ideal to evaluate this method on modern formulas such as Barrett Universal II or Holladay 2, the absence of published equations for these formulas prevents such a study. As such, we studied the application of the machine learning predicted postoperative ACD in four existing formulas whose mathematical equations were published. Although it awaits to be further validated, similar results can likely be transferred to other refraction prediction methods, since many modern IOL power formulas use predicted postoperative ACD as an intermediate step for predicting postoperative refraction.

In summary, the results of this study demonstrate that a machine learning method for postoperative ACD prediction based on postoperative optical biometry can be incorporated into a variety of existing IOL power formulas to improve their accuracy in refraction prediction.

## Data Availability

Data are not publicly available.

## ACKNOWLEDGMENTS

None

## CONTRIBUTIONS

TL: data analysis, programming, and writing of the manuscript; JDS: data collection; NN: data collection, guidance on method development, and writing of the manuscript

## FUNDING

This work was supported by the Lighthouse Guild, New York, NY (JDS) and National Eye Institute, Bethesda, MD, 1R01EY026641-01A1 (JDS).

## COMPETING INTERESTS

None declared

## DATA AVAILABILITY STATEMENT

Data are not publicly available

## Notes

### Competing Interest Statement

The authors have declared no competing interest.

### Author Declarations

This study has been approved by the IRB at the University of Michigan

