## Supplementary materials for "AI-Powered Effective Lens Position Prediction Improves the Accuracy of Existing Lens Formulas"

| Formula | Constant | Optimized Constants | Mean Error |
|---|---|---|---|
| Haigis | a0 (a1 = 0.234, a2 = 0.217) | -0.736 | 0.000319 |
| Hoffer Q | ACD | 5.726 | 0.000157 |
| Holladay1 | Surgeon factor | 1.867 | 0.000121 |
| SRK/T | A constant | 119.091 | -0.000204 |

**Table S1** The optimized A constants and the corresponding mean error for the original formulas in the training dataset. The A constants were optimized so that the absolute value of the mean error was minimized. The mean errors were calculated after excluding the outliers (see main text). The mean errors in the training set were rounded to three significant figures.

| Dataset | Variables | Holladay1 | SRK/T | Hoffer Q | Haigis |
|---|---|---|---|---|---|
| Training dataset | $ELP_F$ | 4.00 ± 0.41 | 5.82 ± 0.50 | 5.86 ± 0.38 | 5.28 ± 0.36 |
| | $ELP_{BC}$ | 4.08 ± 0.92 | 5.85 ± 0.83 | 5.97 ± 1.00 | 5.32 ± 0.76 |
| | $ELP_{ML}$ | 4.67 ± 0.27 | | | |
| Testing dataset | $ELP_F$ | 4.00 ± 0.39 | 5.83 ± 0.47 | 5.87 ± 0.38 | 5.28 ± 0.36 |
| | $ELP_{BC}$ | 4.06 ± 0.72 | 5.84 ± 0.65 | 5.95 ± 0.80 | 5.31 ± 0.61 |
| | $ELP_{ML}$ | 4.68 ± 0.26 | | | |

**Table S2** The mean ± standard deviation (SD) for $ELP_F$, $ELP_{ML}$, and $ELP_{BC}$ in the training and testing dataset when $ELP_F' = ELP_F$. The $ELP_{BC}$ and $ELP_F$ were calculated using the corresponding formula with the optimized A-constants therefore their values vary with different formulas. The values of $ELP_{ML}$ only depend on the values of the preoperative biometry. The outliers were not removed when the above summary statistics were calculated. All values were rounded to two decimal places.

| Methods | Holladay1 | SRK/T | Hoffer Q | Haigis |
|---|---|---|---|---|
| Formula LR | $c_1$ = 1.27<br>$c_3$ = -1.01 | $c_1$ = 0.76<br>$c_3$ = -1.67 | $c_1$ = 1.44<br>$c_3$ = -26.65 | $c_1$ = 1.00<br>$c_3$ = -5.88 |
| ML LR | $c_2$ = 1.65<br>$c_3$ = -3.80 | $c_2$ = 1.31<br>$c_3$ = -0.33 | $c_2$ = 1.62<br>$c_3$ = -1.70 | $c_2$ = 1.24<br>$c_3$ = -0.53 |
| Formula & ML LR | $c_1$ = 0.98<br>$c_2$ = 0.61<br>$c_3$ = -2.77 | $c_1$ = 0.47<br>$c_2$ = 0.79<br>$c_3$ = -2.55 | $c_1$ = 1.09<br>$c_2$ = 0.65<br>$c_3$ = -6.90 | $c_1$ = 0.58<br>$c_2$ = 0.68<br>$c_3$ = -4.44 |

**Table S3** The coefficients ($c_1$ and $c_2$) and the intercept $c_3$ for the linear regression model established based on the training dataset. All values were rounded to two decimal places.

| Dataset | Method | Holladay1 | SRK/T | Hoffer Q | Haigis |
|---|---|---|---|---|---|
| Training dataset | Formula LR | 4.06 ± 0.52 | 5.80 ± 0.38 | 5.90 ± 0.55 | 5.28 ± 0.36 |
| | ML LR | 3.92 ± 0.44 | 5.78 ± 0.35 | 5.86 ± 0.43 | 5.26 ± 0.33 |
| | Formula & ML LR | 3.98 ± 0.53 | 5.80 ± 0.40 | 5.89 ± 0.55 | 5.27 ± 0.37 |
| Testing dataset | Formula LR | 4.04 ± 0.46 | 5.80 ± 0.31 | 5.91 ± 0.53 | 5.28 ± 0.34 |
| | ML LR | 3.90 ± 0.39 | 5.79 ± 0.32 | 5.87 ± 0.39 | 5.27 ± 0.30 |
| | Formula & ML LR | 3.97 ± 0.46 | 5.79 ± 0.34 | 5.89 ± 0.49 | 5.28 ± 0.34 |

**Table S4** The mean ± standard deviation (SD) for $ELP_F'$ in the training and testing dataset. The $ELP_{BC}$ and $ELP_F$ were calculated using the corresponding formula with the optimized A-constants therefore their values vary with different formulas. The values of $ELP_{ML}$ only depend on the values of the preoperative biometry. The outliers were not removed when the above summary statistics were calculated.

| Methods | Holladay1 | SRK/T | Hoffer Q | Haigis |
|---|---|---|---|---|
| Original | -0.020 ± 0.513 | -0.008 ± 0.528 | -0.020 ± 0.529 | -0.025 ± 0.496 |
| Formula LR | 0.008 ± 0.517 | -0.003 ± 0.522 | -0.017 ± 0.492 | -0.018 ± 0.496 |
| ML LR | 0.057 ± 0.563 | 0.001 ± 0.525 | 0.007 ± 0.589 | -0.001 ± 0.523 |
| Formula & ML LR | 0.009 ± 0.500 | -0.008 ± 0.490 | -0.016 ± 0.475 | -0.014 ± 0.484 |

**Table S5** The mean error (ME) ± standard deviation (SD) of alternative linear models in the testing set. All values were rounded to three decimal places.

| **Methods** | **Holladay1** | **SRK/T** | **Hoffer Q** | **Haigis** |
|---|---|---|---|---|
| Original | 0.299 | 0.307 | 0.330 | 0.283 |
| Formula LR | 0.305 | 0.310 | 0.293 | 0.283 |
| ML LR | 0.351 | 0.308 | 0.366 | 0.304 |
| Formula & ML LR | 0.290 | 0.273 | 0.268 | 0.263 |

**Table S6** The median absolute error (MedAE) of alternative linear models in the testing set. All values were rounded to three decimal places.

| **Statistic** | **Holladay1** | **SRK/T** | **Hoffer Q** | **Haigis** |
|---|---|---|---|---|
| Friedman chi-square test statistic | 37.29 | 39.13 | 117.42 | 37.25 |
| p-value | 4.00e-08 | 1.63e-08 | 2.78e-25 | 4.07e-08 |

**Table S7** The Friedman test statistic and the p-values for comparing the testing set results of different methods. All Friedman statistics were rounded to two decimal places. All p-values were rounded to three significant figures.

| **Formula** | **Methods** | ML LR | Formula LR | Formula & ML LR |
|---|---|---|---|---|
| Haigis | Formula LR | **1.7E-01** | **/** | **/** |
| | Formula & ML LR | 1.8E-11 | 2.6E-03 | / |
| | Original | **1.7E-01** | **1.0E+00** | 2.7E-03 |
| Hoffer Q | Formula LR | 1.5E-10 | / | / |
| | Formula & ML LR | 1.7E-26 | 3.6E-05 | / |
| | Original | 4.1E-05 | 1.5E-10 | 5.1E-17 |
| Holladay 1 | Formula LR | 1.5E-04 | / | / |
| | Formula & ML LR | 4.4E-12 | 3.0E-04 | / |
| | Original | 1.4E-05 | **1.0E+00** | 9.9E-02 |
| SRK/T | Formula LR | 1.0E+00 | / | / |
| | Formula & ML LR | 1.7E-12 | 7.0E-06 | / |
| | Original | **1.0E+00** | **1.0E+00** | 1.1E-05 |

**Table S8** The post hoc test results of four existing formulas for comparing the testing set performance of different methods. The insignificant p-values (p≥0.05) were highlighted in bold.

| **Method** | **Formulas** | **Short AL** ($AL < 22mm$) **n=28** | **Medium AL** ($22mm \le AL \le 26mm$) **n=832** | **Long AL** ($AL > 26mm$) **n=100** |
|---|---|---|---|---|
| Original | Haigis | 0.321 ± 0.234 | 0.373 ± 0.332 | 0.383 ± 0.315 |
| | Hoffer Q | 0.524 ± 0.295 | 0.396 ± 0.335 | 0.480 ± 0.350 |
| | Holladay1 | 0.397 ± 0.224 | 0.364 ± 0.322 | 0.541 ± 0.464 |
| | SRK/T | 0.438 ± 0.236 | 0.386 ± 0.337 | 0.452 ± 0.465 |
| Formula & ML LR | Haigis | 0.330 ± 0.285 (-2.8%) | 0.353 ± 0.331 (5.5%) | 0.394 ± 0.319 (-2.8%) |
| | Hoffer Q | 0.338 ± 0.264 (35.5%) | 0.344 ± 0.318 (13.1%) | 0.420 ± 0.338 (12.5%) |
| | Holladay1 | 0.392 ± 0.257 (1.3%) | 0.356 ± 0.320 (2.2%) | 0.486 ± 0.445 (10.3%) |
| | SRK/T | 0.375 ± 0.284 (14.3%) | 0.351 ± 0.324 (9.0%) | 0.438 ± 0.391 (3.2%) |

**Table S9** The mean absolute error (MAE) ± standard deviation in the testing set for patients with short, medium, and long axial length, where $ELP_F' = c_1 \cdot ELP_F + c_2 \cdot ELP_{ML} + c_3$. All MAE and SD were rounded to three decimal places. For $ELP_F' = c_1 \cdot ELP_F + c_2 \cdot ELP_{ML} + c_3$, the percentage reduction in MAE compared to $ELP_F' = ELP_F$ were also shown. The percentage reduction was calculated as $\frac{MAE\ of\ the\ original\ formulas - MAE\ of\ the\ formulas\ using\ adjusted\ ELP_F}{MAE\ of\ the\ original\ formulas} \cdot 100\%$ and was rounded to one decimal place.

### Calculation of $ELP_{BC}$

As described in the main text, the postoperative refraction was predicted using a function of $ELP_F$ and preoperative biometry: $predicted\ refraction = f_1(ELP_F, biometry)$. Here we define $ELP_{BC}$ as follows: when $ELP_F = ELP_{BC}$, $f_1(ELP_{BC}, biometry) - true\ refraction = 0$ holds for all cases. In other words, when the ELP estimation equals $ELP_{BC}$, the refraction prediction error equals zero for all cases. Based on the above definition, the value of $ELP_{BC}$ can be found by solving for the $x$ in the equation $f_1(x, biometry) - true\ refraction = 0$, where $biometry$and $true\ refraction$ are known. For a given case, there were always no more than two roots for the above function because of the quadratic nature of the formulas. When there were two roots, the smaller root was taken as $ELP_{BC}$ because of two main reasons: (1) the greater root was usually >50, which was not within a physiologically meaningful range for ELP; (2) practically when the larger roots were used as $ELP_{BC}$, the $R^2$ in the training set was significantly lower than that obtained with the smaller root (data are not shown). The function $f_1(x, biometry) - true\ refraction = 0$ was solved programmatically using scipy.optimize.fsolve (scipy 1.2.1) in Python 3.7.3.

### A-Constant Optimization

When $ELP_F$ was not replaced with a modified value $ELP'_F$, the A-constants of the formulas were optimized in the standard way: first, compute the mean refraction prediction error when the A-constant takes different values, then, the A-constant that gives the smallest absolute mean error is the most optimal A-constant.

When $ELP_F = ELP'_F$, the A-constants were optimized based on the same concept, the value of $ELP'_F$ changes with the values of the A-constant. The pseudo-code for the A-constant optimization process is shown below. The value of $ELP_{ML}$ does not change with the A-constant. The value of $ELP_F$ and $ELP_{BC}$ changes with the A-constants.

**Algorithm 1**: A-constant optimization when $ELP_F = ELP'_F$

$ELP_{ML}$ ← compute $ELP_{ML}$ using the machine learning model
**FOR** $a$ **IN** A-constant search space
    $ELP_F$ ← compute $ELP_F$ based on the formula with $a$ as the A constant
    $ELP_{BC}$ ← compute $ELP_{BC}$ based on the formula with $a$ as the A constant
    coefficients $c_1$, $c_2$, and $c_3$ ← model $ELP_{BC}$as a linear function of $ELP_{ML}$and/or $ELP_F$.
    $ELP'_F \leftarrow c_1 \cdot ELP_F + c_2 \cdot ELP_{ML} + c_3$
    predicted refraction ← compute the predicted refraction based on $a$ and $ELP'_F$
    mean error ←compute the mean error based on the predicted refraction and the true refraction
**END FOR**
The most optimal A-constant← the A-constant that gives the smallest absolute mean error
